# STUDY TO ASSESS ASSOCIATION OF C-REACTIVE PROTEIN WITH NEPHROPATHY IN PEOPLE LIVING WITH TYPE 2 DIABETES MELLITUS

**DOI:** 10.1101/2020.09.23.20200519

**Authors:** Kaushik Mondal, Dattatreya Mukherjee

## Abstract

**Introduction:** **Diabetes mellitus** (DM) is a metabolic disorder with inappropriate hyperglycemia either due to an absolute or relative deficiency of insulin secretion or reduction in the biologic effectiveness of insulin or both. An inflammatory basis for Diabetes & its complications has attracted interest. Among several markers of inflammation, **C reactive protein** (CRP) is found to be significant in people with diabetes. Diabetic nephropathy may be associated with abnormally high levels of CRP.

**Materials & Methods:** This study was performed for 1 year over 100 type 2 diabetic patients (n=100) aged 30-70 years in Calcutta National Medical College & Hospital. Necessary clinical & Diabetes related laboratory investigations were done as the routine procedure.

**Results:** Among 100 study population, all macro-albuminuria patients were found to have high CRP. 63% of Stage 3 chronic kidney disease (CKD) patients were having high CRP in our study. High CRP values were also found among patients with higher spot urine ACR (329.50±259.40 mg/gram).(P<0.05)

**Conclusion:** CRP is a marker which increased in inflammatory reactions. Low grade inflammation as indicated by high CRP was an important predictor of diabetic nephropathy.

## Introduction

Diabetes mellitus (DM) is a metabolic disorder with inappropriate hyperglycemia either due to an absolute or relative deficiency of insulin secretion from beta cells of langerhans of endocrine part of pancreas or reduction in the biologic effectiveness of insulin or both or the outer receptors are malfunctioning. It is one of the most common non-communicable diseases worldwide and is a growing concern. Diabetes prevalence had increased by 64% across India over the last quarter century according to November 2017 report by Indian Council for Medical Research, Institute for Health Metrics and Evaluation, both research institutes and the Public Health Foundation of India, an advocacy. Genetics, Aging, Obesity, Lifestyles & many other factors have contributions to the development of Diabetes and its complications. Nevertheless an inflammatory basis for Diabetes and its complications has attracted interest. Among several markers of inflammation, hs–CRP is found to be significant in people with diabetes. CRP, a pentameric protein produced by the liver has emerged as the ‘golden marker for inflammation’. Proteinuria is a marker of vascular endothelial damage. Additionally, it is reported that high serum levels of CRP is a novel cardiovascular risk factor that impairs endothelial function. So diabetic nephropathy may be associated with abnormally high levels of CRP.

Diabetic nephropathy is a leading cause of CKD, end stage renal disease(ESRD). It is diagnosed based upon the detection of proteinuria in diabetic patients in absence of other obvious cause such as infection^1^.

Definition of diabetic nephropathy by albuminuria^2^:

In our study, we used spot urinary albumin-Creatinine ratio for detection of albuminuria. We tested spot urine sample after excluding conditions that could transiently increase albumin excretion and same test was repeated 2 times at three months interval. If two out of three tests were positive, we considered the people having diabetic nephropathy.

Non-diabetes related conditions that might increase albuminuria are urinary tract infection(UTI), haematuria(RBC>5 RBC/hpf), prostate disease, heart failure, febrile illness, severe hypertension and vigorous exercise.

Microalbuminuria and macroalbuminuria may be present when patient is diagnosed of having type 2 DM first reflecting its long asymptomatic period. To institute effective therapy at an early stage, albuminuria should be detected. But some individuals with type 1 and type 2 DM have a decline glomerular filtration rate (GFR) in the absence of albuminuria. So measurement of the serum Creatinine & estimation of GFR should also be performed.

For patients with type 2 diabetes mellitus in NHANES III (THIRD NATIONAL HEALTH AND NUTRITION EXAMINATION SURVEY; n=1197), low GFR (60 ml/min/1.73 m^2^) was present in 30% of patients in absence of micro or macroalbuminuria and retinopathy^3^.

GFR can be measured by specific techniques, example-lnulin clearance. But the recommended equation by the National Kidney Foundation is that of the MDRD (Modified Diet In Renal Disease):estimated GFR (ml/min/1.73m^2^)=186* (Serum creatinine in mg/dl)^-1.154 *^(age in years)^-0.203^. In female, it should be multiplied by 0.742.

Compared to MDRD formula, Inulin clearance slightly overestimates the glomerular functions. In early stage of renal disease, Inulin clearance may remain normal due to hyperfiltration in the remaining nephrons^4^.

GFR can also be accurately measured using radioactive substances, in particular Chromium-51 and Technetium-99m. These come close to ideal properties of Inulin but can be measured more practically with only a few blood or urine samples^5^. However we did not use such sophisticated procedures considering the financial burden to the patients.

### Stage of chronic kidney disease based on e GFR

But one point should also be noted that glomerular hyperperfusion and renal hypertrophy occur in the first years after the onset of DM and are associated with increase in GFR.

We did not include haemodialysis patients in our study as it was seen that there were some dialysis related alterations in the immune and host defence system. Moreover the type of dialysis membrane had been suggested to play a role. So haemodialysis might exert an impact on C-Reactive protein level.

Creatinine clearance using CockCroft-Gault equation is less accurate. Creatinine clearance (ml/min)= {(140-age in years)* weight in kg}/(72* serum creatinine in mg/dl). It should be multiplied by 0.85 in females. Normal creatinine clearance for healthy men is 97-137 ml/min and for healthy women is 88-128 ml/min^6^.

Nephropathy may be present due to conditions other than diabetes mellitus in people living with type 2 Diabetes. To establish diabetic nephropathy, some points should be kept in mind.

a. Duration of diabetes and glycemic status
b. Presence of hypertension
c. Associated retinopathy
d. Absence of haematuria and RBC (red blood cells) casts
e. Absence of strong family history of kidney disease due to etiology other than diabetes
f. Absence of rapid rise of creatinine.

We have only included patients clinically and biochemically suspected to have diabetic kidney disease.

Normal blood value of CRP is considered as <4 mg/l according to laboratory reference range of our hospital.

## Objectives

1. To correlate CRP level in relation to normo albuminuria, micro albuminuria and macro albuminuria.
2. To correlate CRP protein levels with diabetic nephropathy based on estimated glomerular filtration rate.
3. To assess the influences of various clinical and biochemical factors on development of diabetic nephropathy.

## Materials & Methods

Total 100 type 2 diabetic patients (n=100) aged 30-70 years were studied for 1 year in Calcutta National Medical College & Hospital.

a. <u>Inclusion criteria</u>-Previously diagnosed or recently diagnosed type 2 Diabetes mellitus patients (age 30-70 years) were included in this study.
b. <u>Exclusion criteria</u>: Patient with a history of urinary tract infections/ pyelonephritis, nephrolithiasis (kidney or bladder stone), catheterization, severe uncontrolled hypertension(HTN) (> 160/100 mm Hg), sickle cell anaemia, patients on haemodialysis, cancer (prostate, bladder and kidney), benign prostatic hypertrophy, polycystic kidney disease(PKD), prolonged use of analgesics, any recent infection which was caused by an infectious etiology and any type of surgery were excluded from the current study.

Inflammatory processs like systemic lupus erythematosus, inflammatory bowel disease, rheumatoid arthritis, giant cell arteritis, osteomyelitis, rheumatic fever and tuberculosis were also excluded.

▪ Clinical examinations & necessary laboratory investigations were done including blood for CRP, HbA1C, lipid profile, Creatinine and spot urine ACR. **CRP was measured by turbidimetry method**.

All statistical analyses were performed using Graph Pad Instat software (Version 3.10, 32 bit for Windows). The chi-square test and the Fisher exact test were used to evaluate the differences in proportions between the two groups. One way *ANOVA test* was used to compare groups for continuous variables. *Tukey Kramer Multiple Comparison test* was done after ANOVA if results were significant overall to find out which specific group’s mean was different. P-value <0.05 was taken as the level of significance.

## Results

Table 3 & Figure 1 are showing distributions of CRP among patients with proteinuria in our study population. Of 17 patients having normoalbuminuria, 16 patients were having normal CRP (94.12%). Of 28 patients having macroalbuminuria, all were having raised CRP (100%). However raised CRP was found in 26 patients out of 55 patients having microalbuminuria (47.3%). (P<0.05)

**TABLE 1:**

| Urine specimen | Microalbuminuria | Macroalbuminuria |
| --- | --- | --- |
| 24 hour collection | 30-299 mg/day | >=300 mg/day |
| Albumin concentration | 20-300 mg/l | >300 mg/l |
| Spot albumin- creatinine ratio | 30-300 mg/gram | >300mg/gram |

**TABLE 2:**

| Stage | e GFR (ml/min/1.73 m<sup>2</sup>) |
| --- | --- |
| 1 | >=90 |
| 2 | 60-89 |
| 3A | 45-59 |
| 3B | 30-44 |
| 4 | 15-29 |
| 5 | <15 |

**TABLE 3.** DISTRIBUTION OF CRP ACROSS THE RANGE OF PROTEINURIA.

**DISTRIBUTION OF CRP ACROSS THE RANGE OF PROTEINURIA**
|  | **CRP (<4**<br>**mg/dl)**<br>**(n=44)** | **CRP (>= 4**<br>**mg/dl)**<br>**(n=56)** |
| --- | --- | --- |
| **Normoalbuminuria** | 16 | 1 |
| **Microalbuminuria** | 29 | 26 |
| **Macroalbuminuria** | 0 | 28 |

**FIGURE 1.**
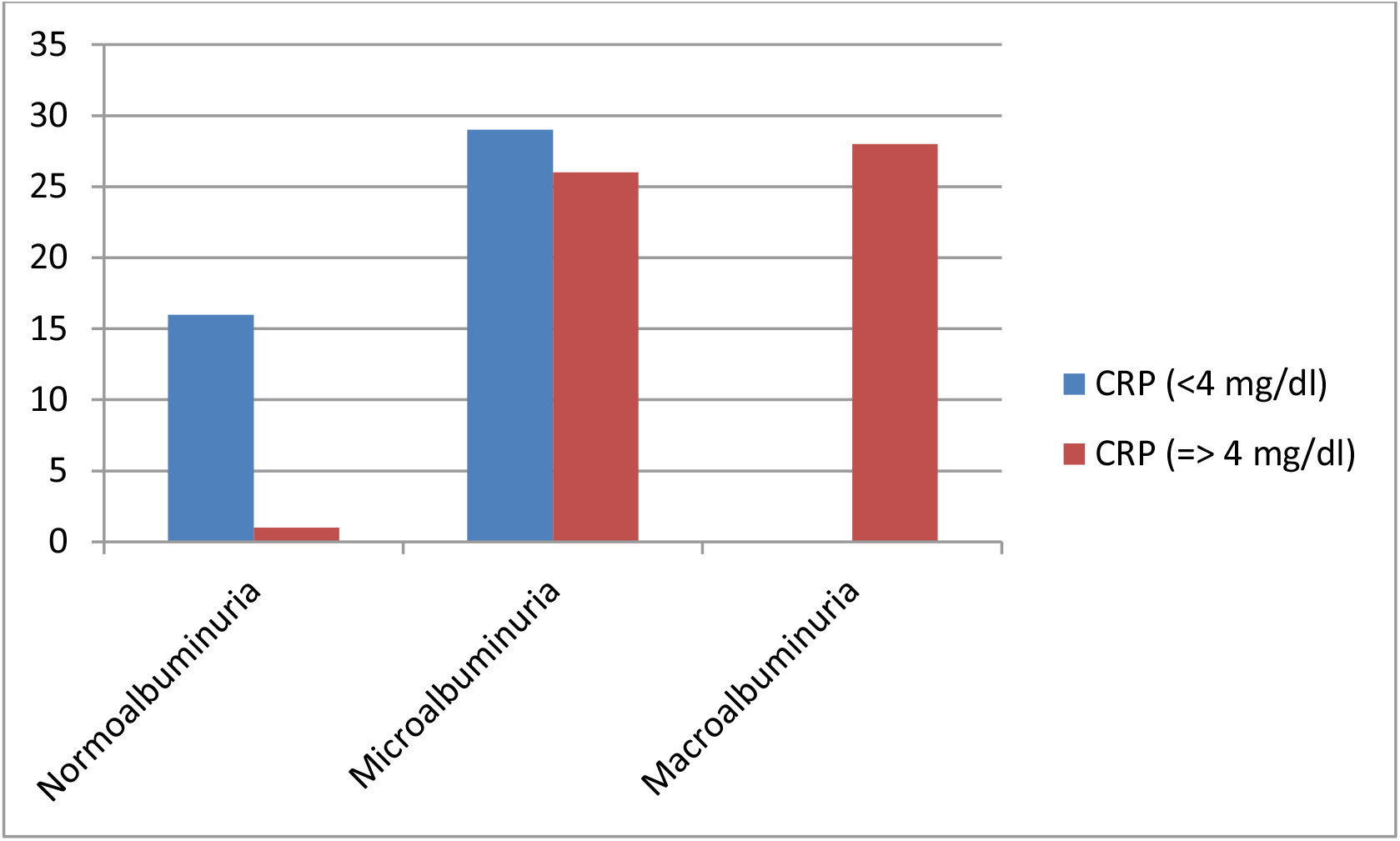
Bar diagram showing distribution of CRP across the range of proteinuria.

Table 4 & Figure 2 are showing 26.3% of Stage 1 chronic kidney disease patients were having high CRP. 63% of Stage 2 chronic kidney disease patients were having high CRP. 63% of Stage 3 chronic kidney disease patients were having high CRP. (P<0.05) Association of eGFR with CRP was significant.

**TABLE 4.** CRP DISTRIBUTION IN RELATION TO STAGES OF CHRONIC KIDNEY DISEASE BASED ON eGFR.

|  | CRP<4 mg/dl<br>(n=44) | CRP>4 mg/dl<br>(n=56) |
| --- | --- | --- |
| Stage 1 | 14 | 5 |
| Stage 2 | 14 | 24 |
| Stage 3 | 16 | 27 |

**FIGURE 2.**
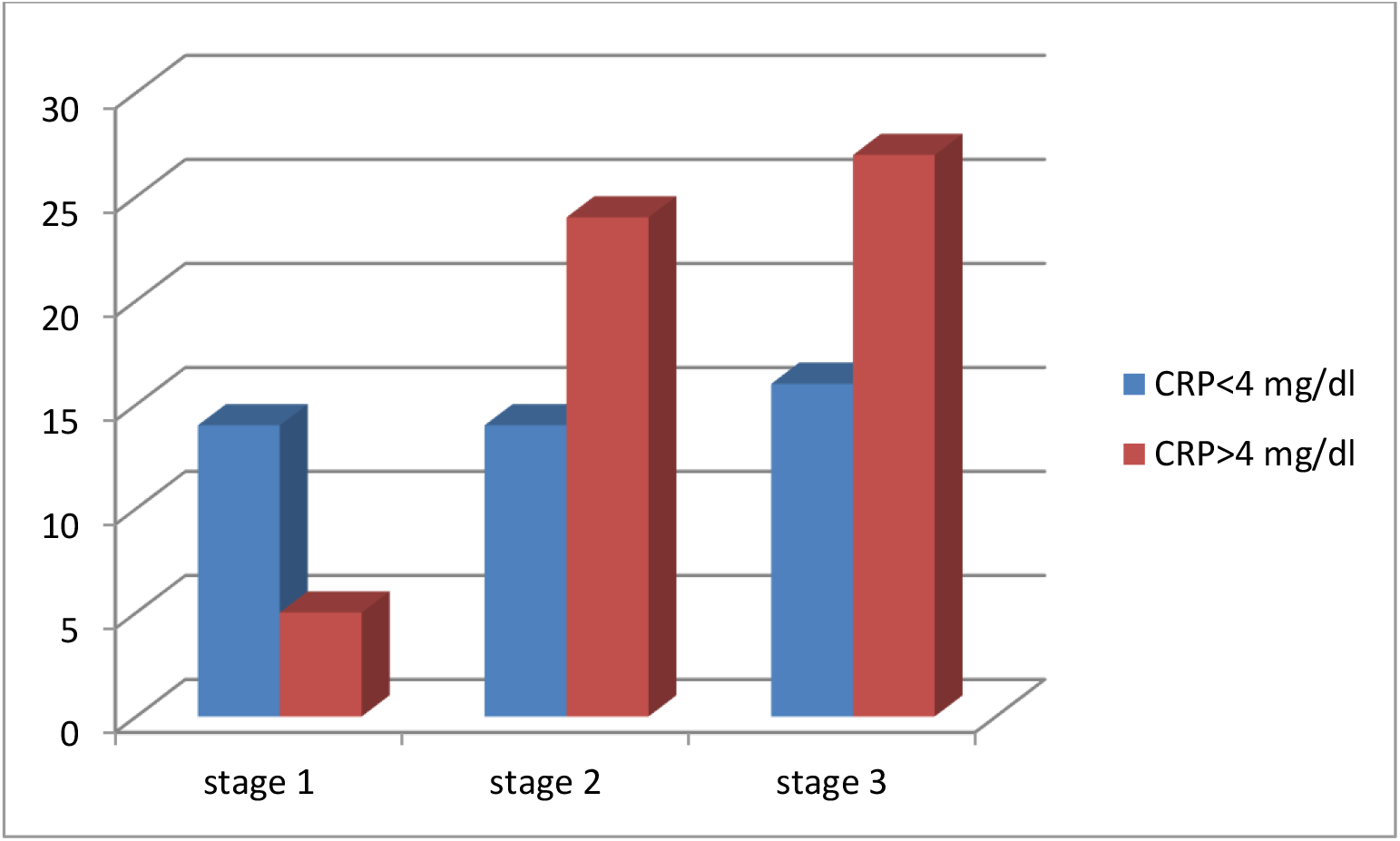
Bar diagram showing CRP distribution in relation to stages of chronic kidney disease based on eGFR.

**FIG 3:**
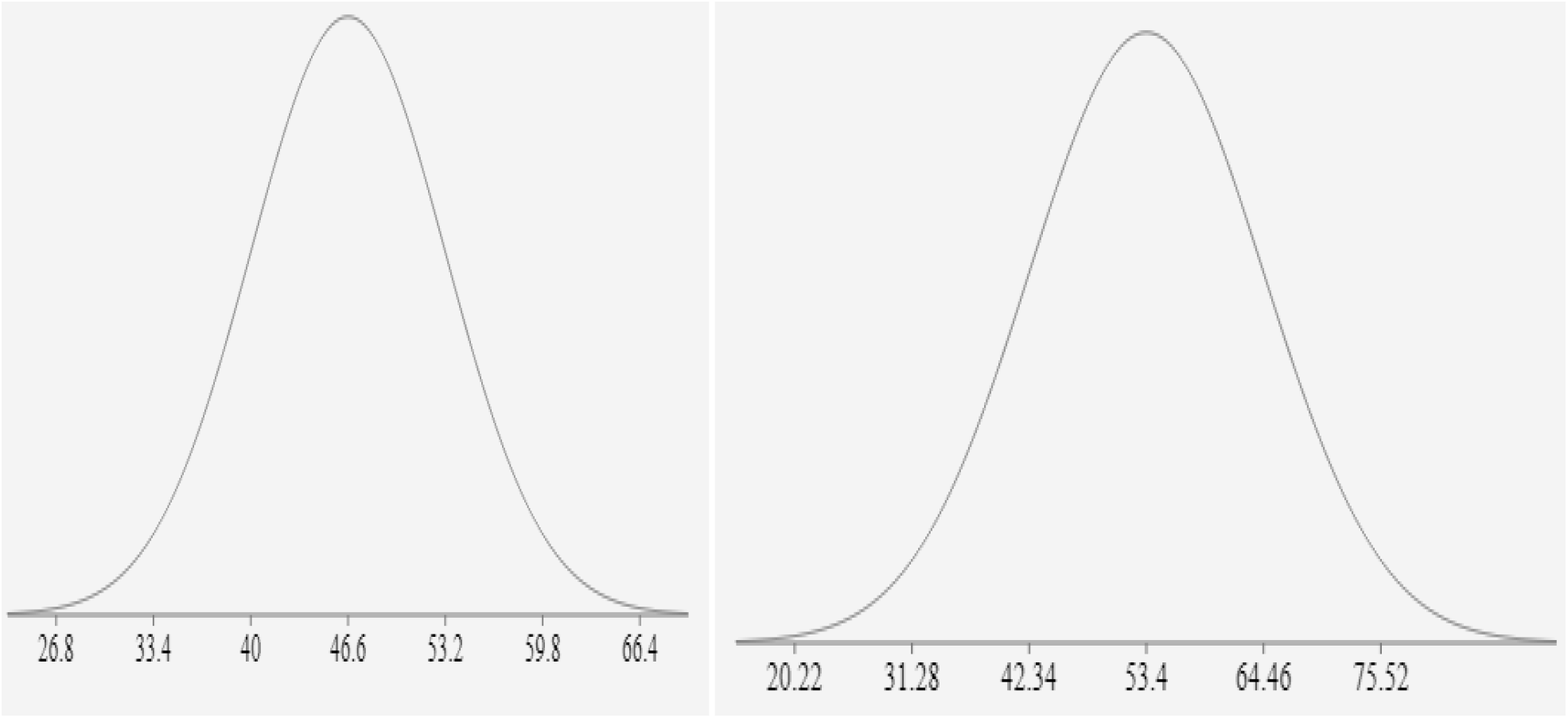
AGE DISTRIBUTION OF Crp<4 and Crp>=4.

**FIG 4.**
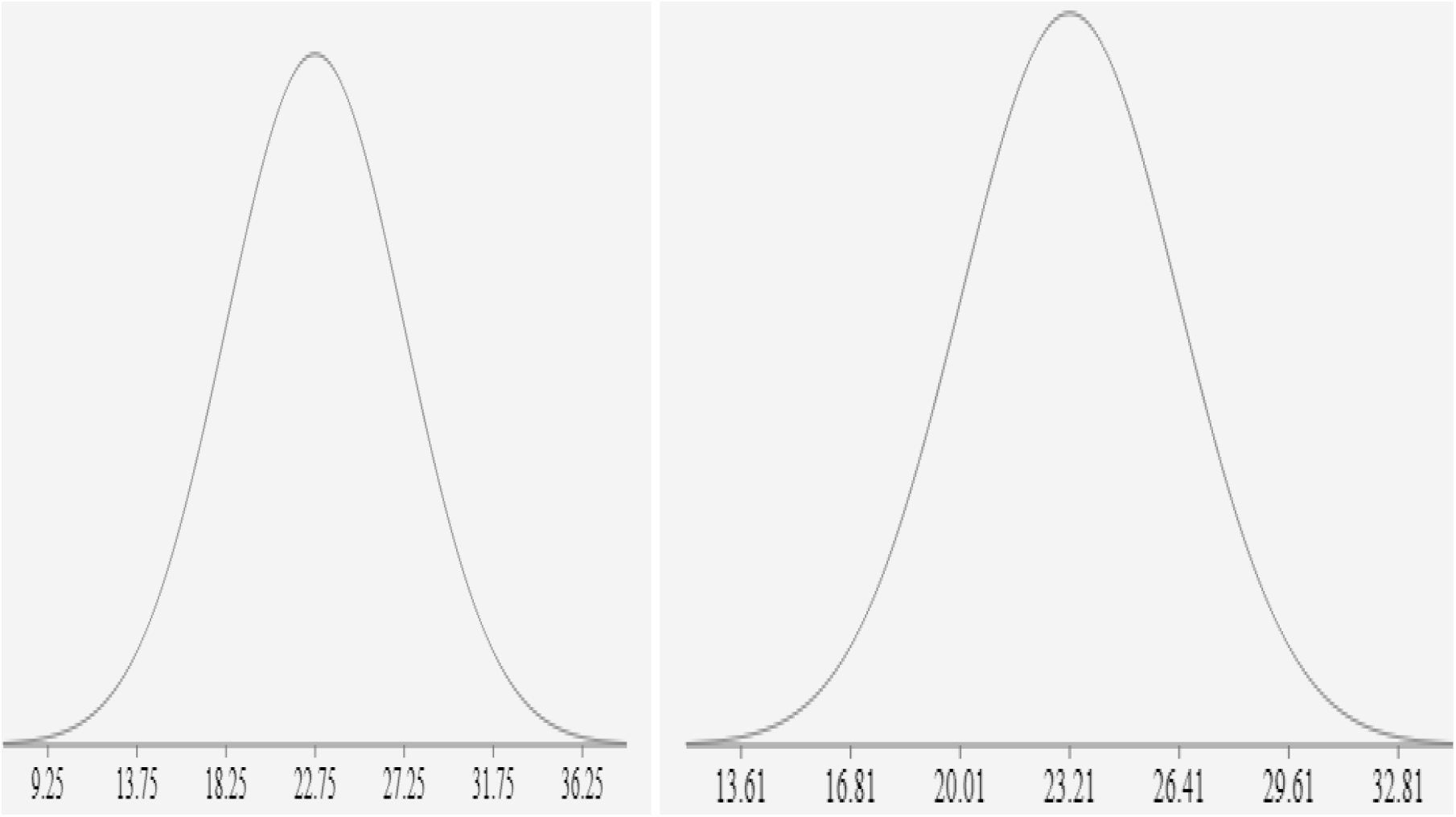
BMI Distribution of CRP<4 and CRP>=4.

**FIG 5.**
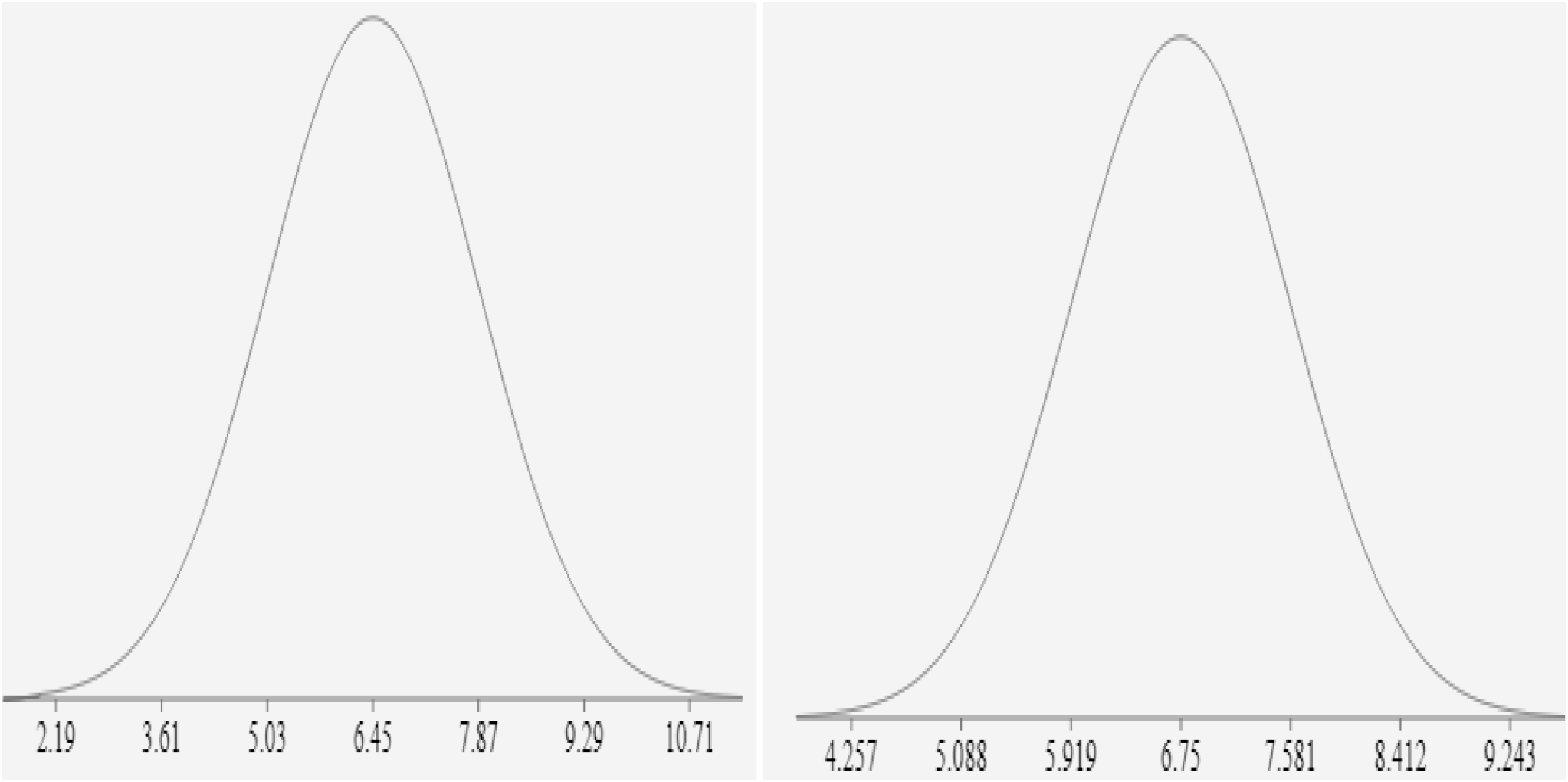
HBA1C Distribution in CRP<4 and CRP>=4 6.

**FIG 6.**
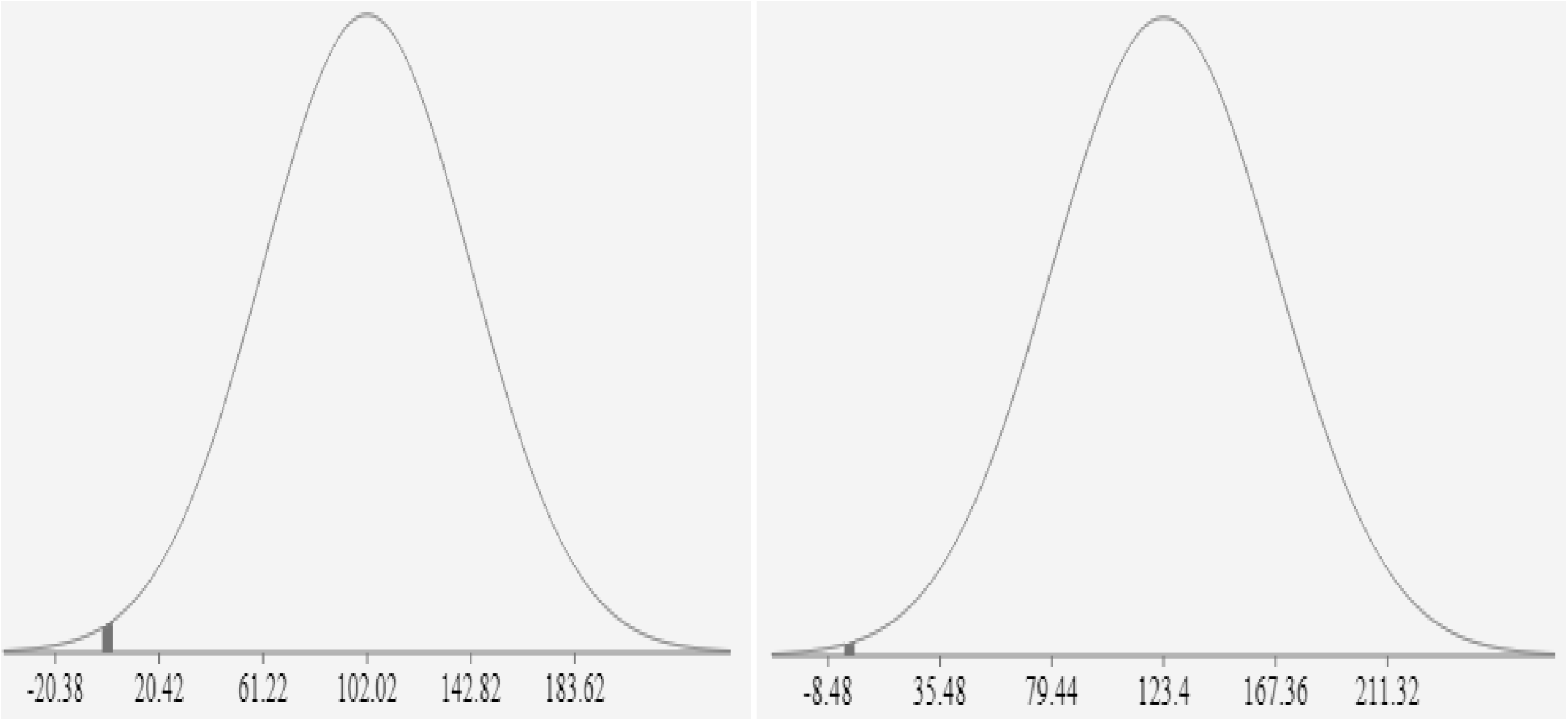
TG distribution in CRP<4 and CRP>=4.

**FIG 7.**
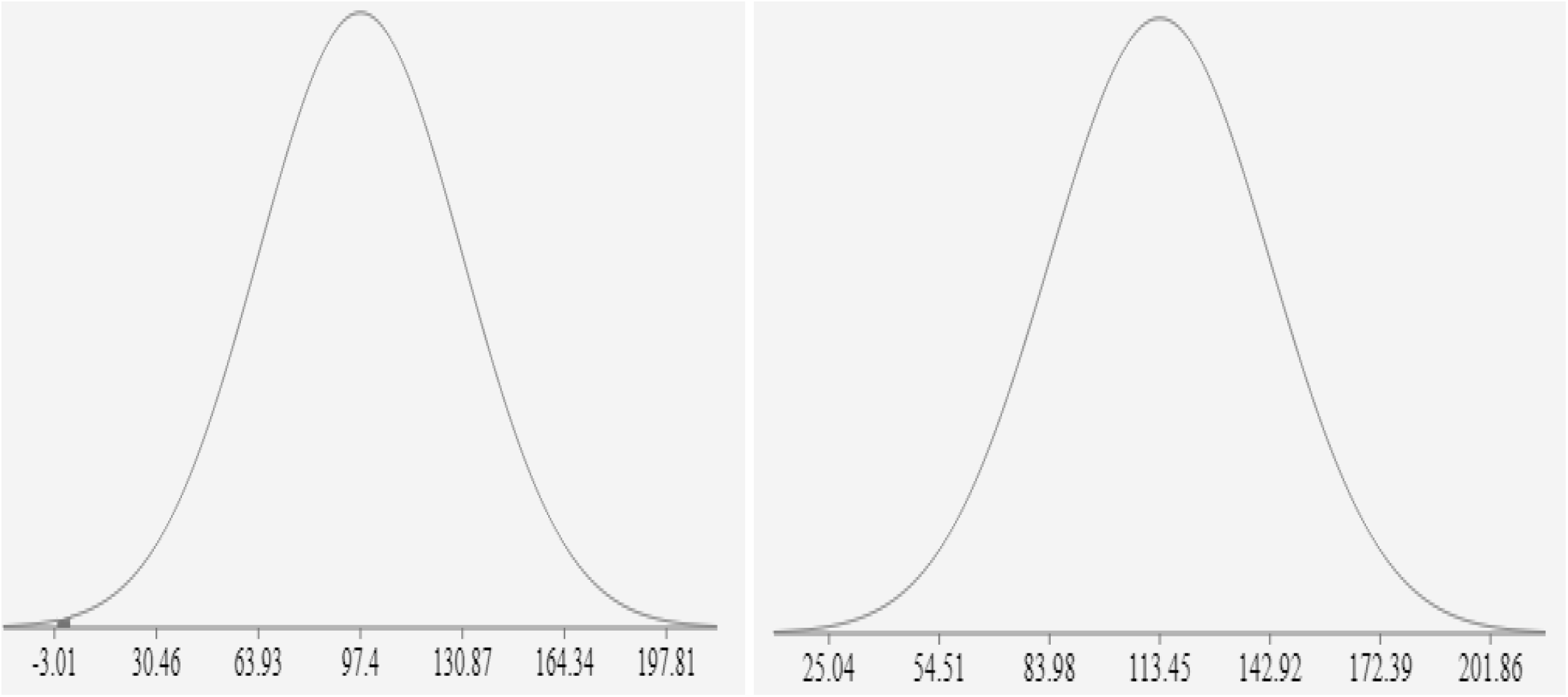
LDL distribution of CRP<4 and CRP>=4.

**FIG 8.**
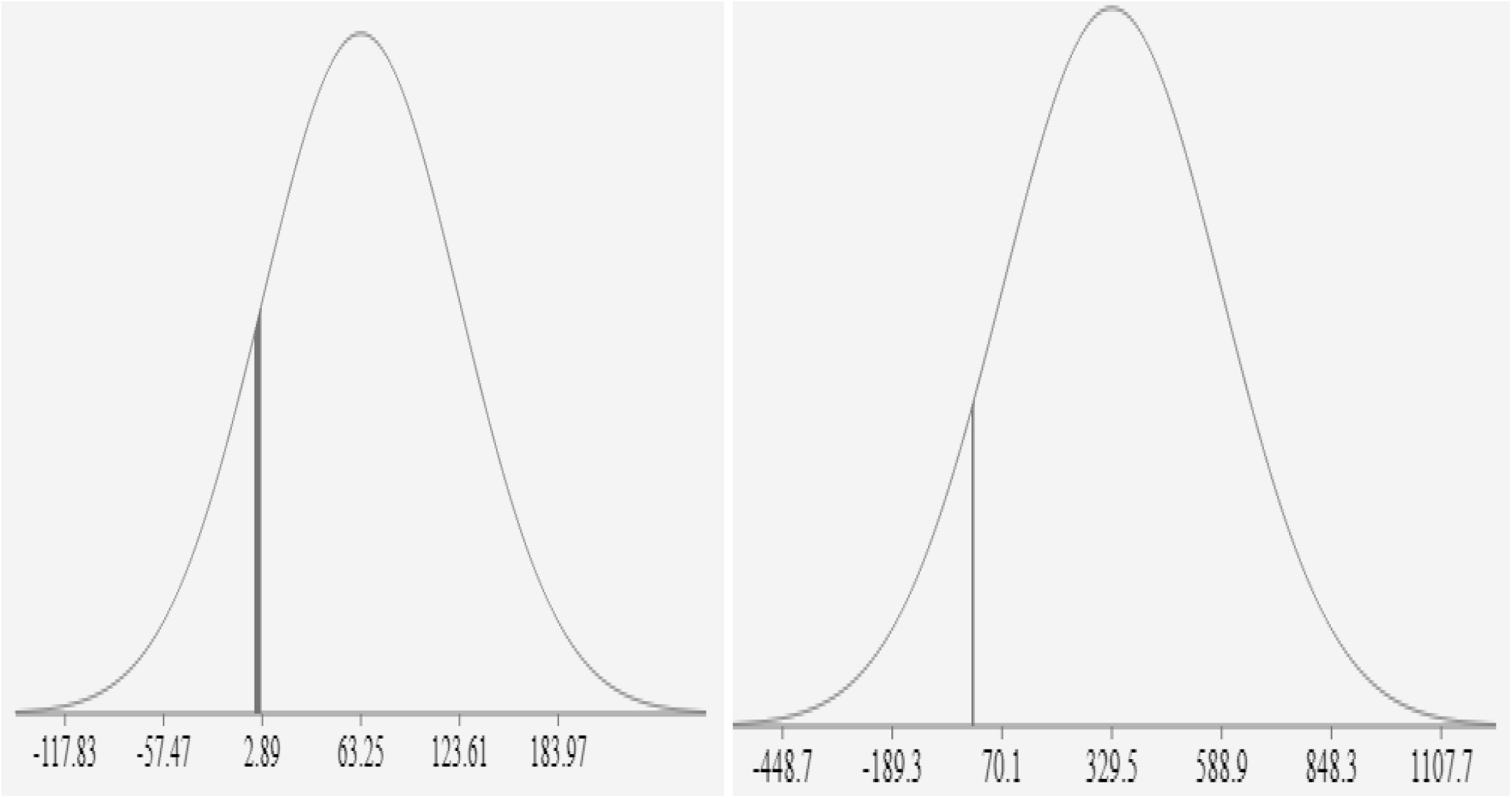
Spot urine ACR distribution in CRP<4 and CRP>=4.

**Fig 9.**
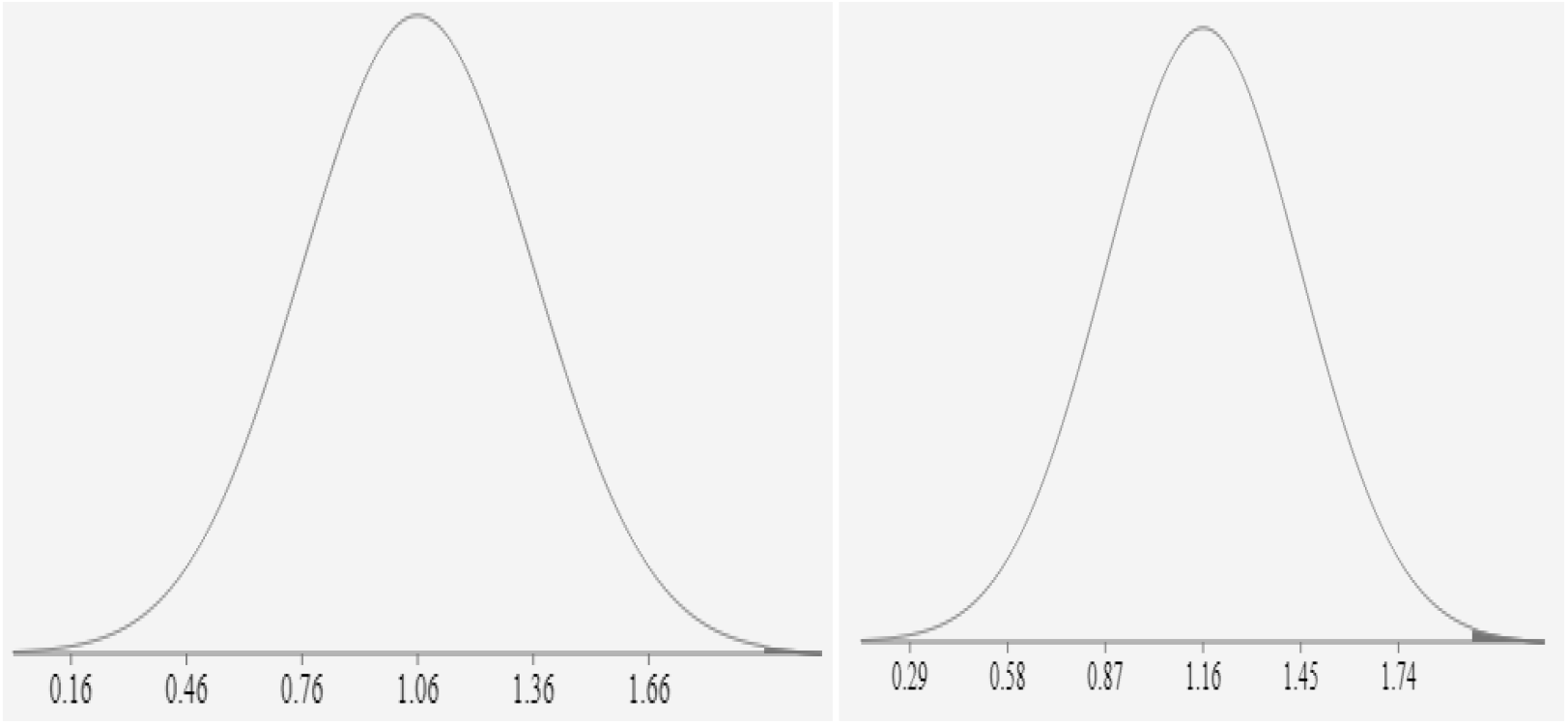
Serum Creatinine Distribution in CRP<4 and CRP>=4.

High CRP values were found in older people (P<0.05) and with higher triglyceride & LDL value (P<0.05). High CRP values were also found among patients with higher spot urine ACR (329.50±259.40 mg/gram) (P<0.05) & with higher creatinine values (P<0.05). However CRP values did not correlate with BMI (P=0.5), HbA1C (P=0.2) (Table 5)

**TABLE 5.** Characteristics Of Diabetic patients with normal and abnormal CRP values :

|  | CRP <4 mg/l | CRP ≥ 4mg/l | P value |
| --- | --- | --- | --- |
|  | (n=44) | (n=56) |  |
| AGE (Year) | 46.6±6.6 | 53.4±11.06 | <0.05 |
| BMI (kg/m<sup>2</sup>) | 22.75± 4.5 | 23.21±3.2 | 0.5 |
| HbA1C (%) | 6.45±1.42 | 6.75±0.831 | 0.2 |
| Triglyceride (TG)(mg/dl) | 102.02± 40.08 | 123.4±43.96 | <0.05 |
| LDL (mg/dl) | 97.4± 33.47 | 113.45± 29.47 | <0.05 |
| Spot Urine ACR (mg/gram) | 63.25±60.36 | 329.50±259.40 | <0.05 |
| **Serum creatinine**<br>**(mg/dl)** | **1.06± 0.3** | **1.16±0.29** | **<0.05** |

The clinical & biochemical characteristics of diabetic patients with proteinuria were represented in Table 6. Patient with macroalbuminuria (spot urine ACR >300 mg/gm) were older & having higher LDL and Serum creatinine and CRP values in comparison to patient with normal urine ACR (<30 mg/gm).

**TABLE 6.** Characteristics of Diabetic Patients With Proteinuria.

|  | NORMOALBUMINURIA | MICROALBUMINURIA | MACROALBUMINURIA | P value |
| --- | --- | --- | --- | --- |
|  | (Spot urine ACR<30mg/gm) | (Spot urine ACR 30-300 mg/gm) | (Spot urine ACR >300 mg/gm) |  |
|  | (n=17) | (n= 55) | (n=28) |  |
| Age( years) | 43.64± 4.91 | 48.69± 9.5 | 58±8.4 | <0.05 |
| BMI (kg/m2) | 23.39± 6.82 | 22.96± 3.54 | 22.86± 1.56 | 0.9 |
| Triglyceride (mg/dl) | 110.4± 46.77 | 113.05± 48.97 | 123.25± 25.88 | 0.5 |
| LDL (mg/dl) | 84.05± 18.78 | 104.44± 30.75 | 123.78± 32.92 | <0.05 |
| HbA1C (%) | 6.44± 1.06 | 6.66± 1.00 | 6.65± 0.67 | 0.6 |
| Serum creatinine (mg/dl) | 0.89± 0.2 | 1.15± 0.3 | 1.19± 0.2 | <0.05 |
| CRP( mg/l) | 1.85±1.5 | 5± 5.09 | 9.17± 4.18 | <0.05 |

Proteinuria was significantly associated with age, serum creatinine, LDL and CRP values overall. But further post hoc analysis (Table 7) showed that age had no significant association while comparing normoalbuminuria with microalbuminuria. The data comparing normoalbuminuria & microalbuminuria which were nearly age matched, HbA1c & triglyceride matched, significant rise of CRP was evident in microalbuminuric patients in comparison to normoalbuminuric patients. So CRP may have significant role in early stages of diabetic nephropathy.

**Table 7.**

|  | AGE | LDL | SERUM CREATININE | CRP |
| --- | --- | --- | --- | --- |
| Normo vs<br>Microalbuminuria | P<0.05 | P<0.05 | P<0.05 | P<0.05 |
| Normo vs<br>Macroalbuminuria | P<0.05 | P<0.05 | P<0.05 | P<0.05 |
| Micro vs<br>Macroalbuminuria | P<0.05 | P<0.05 | **<u>P>0.05</u>** | P<0.05 |

## Discussion

Acute phase proteins are a class of proteins & their plasma concentrations increase or decrease in response to inflammations. Previous studies had demonstrated that inflammation played an important role in the pathogenesis of DM^7-9^.

CRP induces impaired self-regulation of glomerular pressure(GP) and dysfunction of glomerular endothelium^10^. So there is loss of albumin in urine through damaged glomeruli. These observations suggest that low-grade inflammation, reflected by high serum hs-CRP levels, may play a role in the induction of proteinuria^11^.

The study of Stehouwer CD et al in 2002 concluded that in type 2 diabetes, increased urinary albumin excretion, endothelial dysfunction, and chronic inflammation were interrelated processes that would develop in parallel & would progress with time^12^. Chronic inflammation as evidenced by high CRP was significantly associated with duration of diabetes mellitus according to Mojahedi et al^13^.

As proteinuria & CRP are the markers of systemic endothelial dysfunction & preclinical arterial inflammation as well^13,14^, they may have a role to develop other macrovascular complications including cardiovascular diseases^15^. We had also studied serum triglyceride level & serum LDL level in association with CRP. Serum triglyceride is associated with atherosclerosis^16^ & serum LDL is associated with coronary heart disease^17^. There were significant associations between CRP & serum triglycerides as well as between CRP and LDL in our study. Our findings were supported by Manoj Sigdel et al^18^. However Mojahedi et al^13^ found significant association of CRP with serum triglyceride only, not with LDL.

Few previous studies showed higher HbA1C was associated with higher CRP & they had significant association^19^. But we did not find any association between HbA1C & CRP possibly because of the presence of fairly controlled HbA1C in a tertiary hospital with proper follow up.

In some studies, they had found that there was a relationship between proteinuria and age^20-23^. We found similar result too. But there was also an association between CRP & age in our study. It might be the possibility that chronic inflammation, being a background process of diabetic nephropathy which in turn increased with advancement of the age, was leading to rise in CRP. It was very well understood that the levels of hs-CRP significantly associated with age and positively related to insulin resistance as concluded by Safiullah Amanullah et al^24^. Besides Aronson et al. reported that CRP levels among middle-aged people were higher in those with Diabetes mellitus when compared with the healthy subjects^25^.

We did not find any association of proteinuria or hs-CRP with body mass index similar to Mojahedi et al^13^. But it is opposing other studies^26,27^including Stehouwer CD et al^12^.

Serum creatinine in this study was also associated with proteinuria & CRP opposing the finding of Mojahedi et al^13^.

## Conclusions

1. In our present study, we found that low grade inflammation as indicated by high CRP was an important predictor of diabetic nephropathy.
2. It was also found that advanced age in diabetics, dyslipidemia (higher triglycerides and LDL values), higher serum creatinine were also the major correlates with high CRP values.
3. Proteinuria is now widely accepted as an independent risk factor for cardiovascular morbidity and mortality^28,29^. Thus measurement of CRP in diabetic nephropathy can also estimate the cardiovascular risks.
4. As our study population was from a tertiary hospital with fairly controlled HbA1C, we found no significant difference in HbA1C levels across proteinuria levels.

## Limitations of the study

1. Small study population
2. A tertiary hospital based study
3. Study period was short.
4. Mostly lower income group & those belong to rural areas were represented in government hospital, so this study was not the true representative to the society as a whole.
5. Few studies also demonstrated that antidiabetic drugs (Metformin/Pioglitazone) and statins might have effects to reduce inflammation^30^. We could not exclude the patients from our study whom were already on antidiabetic &/or Statin treatment.
6. This was a cross-sectional study. So we could not follow up the study population to assess the further consequences of inflammation and interventions.

## Data Availability

WE HAVE INCLUDED ALL THE DATA FROM THE TEST REPORTS OF THE PATIENT. WE HAVE USED ANNOVA STATISTICAL SOFTWARE FOR STATISTICAL ANALYSIS. ALL THE DATA AND CALCULATIONS ARE CORRECT AND VALID. TESTS ARE DONE FROM VALID LABS.PATIENT CONSENT IS ALSO TAKEN.

## Acknowledgement

We are thankful to the professors of Internal Medicine department and labs of Calcutta National Medical College, West Bengal University of Health Science, India. We are also thankful to the professors of Jinan University for supporting in the research work.

## Author

1. Dr. Kausik Mondal, MBBS, MD (Internal Medicine). Senior Resident (SR) in Jhargram Super specialty hospital, West Bengal, India.
2. Dattatreya Mukherjee, 5^th^ year MBBS student and Researcher in Jinan University, P.R China.

## Ethical Approval

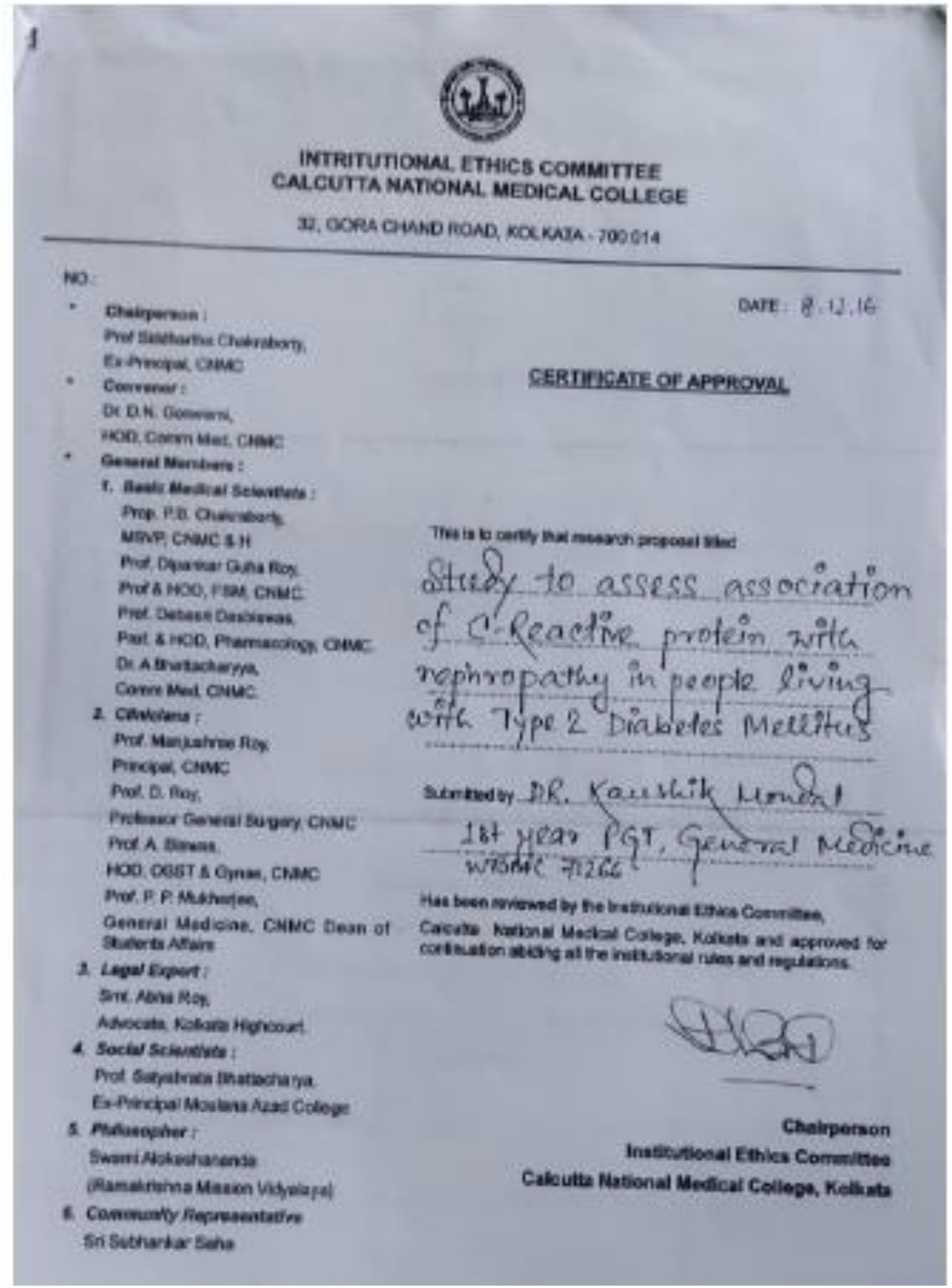

## Bibilograph

1. Rudy Bilous & Richard Donnelly: Handbook of Diabetes; Wiley-Blackwell. 4th edition, page:5

2. Alvin C. Powers: Harrison’s Principles of Internal Medicine; Diabetes Mellitus : Complications, McGraw Hill Education. 19th edition, page-2424–2425

3. Kramer HJ et al, Renal insufficiency in the absence of albuminuria and retinopathy among adults with type 2 Diabetes Mellitus, JAMA 289: 3273–3277, 2003

4. ^GFR (Cockcroft & MDRD) Calculator at medical-calculator.nl –Cockcroft-Gault and MDRD calculator and details about Inulin clearance

5. ^Murray, A. W.., Barnfield, M.C., Waller, M.L., Telford, T., Peters, A. M. (8 may 2013). “Assessment of Glomerular Filtration Rate Measurement with Plasma Sampling: A Technical Review”. Journal of Nuclear Medicine Technology. 41 (2):67–75. Doi:10.2967/jnmt.113.121004

6. Cockcroft, D.W. and M.H. Gault. Prediction of creatinine clearance from serum creatinine. Nephron. 1976. 16(1):31–41.

7. Tsunoda K et al. “Retinopathy and Hypertension affect serum high sensitivity C-reactive protein levels in type 2 Diabertic patients.” J Diabet Complications. 2005;19(3):123–7

8. Kang ES et al. “Relationship of serum high sensitivity C-reactive protein to metabolic syndrome and microvascular complications in type 2 diabetes.” Diabetes Res Clin Pract 2005; 69(2):151–9

9. Thorand B et al. “C-reactive protein as a predictor for incident diabetes mellitus among middle aged men: results from the MONICA Augsburg cohort study. Arch Intern Med. 2003 : 163; 93–9

10. Moreno Ruiz I et al. “Microvascular complications and risk factors in patients with Diabetes.” Endocrinol Nutr. 2011;58 (4) 163–8

11. Yokoyama H et al. “Raised serum sialic acid concentration precedes onset of microalbuminuria in IDDM.” Diabetes care. 1996;19:435–40

12. Stehouwer CD et al. “Increased urinary albumin excretion, endothelial dysfunction, and chronic low-grade inflammation in type 2 diabetes: progressive, interrelated, and independently associated with risk of death.” Diabetes. 2002 Apr;51(4):1157–65

13. Mohammad Javad Mojahedi et al. “Elevated Serum C-Reactive Protein Level and Microalbuminuria in Patients With Type 2 Diabetes Mellitus”. Iranian Journal of Kidney Diseases | Volume 3 | Number 1 | January 2009

14. Torzewski M et al. “C-reactive protein in the arterial intima: role of C-reactive protein receptor dependent monocyte recruitment in atherogenesis.” Arterioscler Thromb Vasc Biol. 2000;20:2094–9

15. Andreas Pfützner et al. “High-Sensitivity C-Reactive Protein Predicts Cardiovascular Risk in Diabetic and Nondiabetic Patients: Effects of Insulin-Sensitizing Treatment with Pioglitazone.” J Diabetes Sci Technol 2010;4(3):706–71

16. Talayero BG et al. “The role of Triglycerides in atherosclerosis.” Curr Cardiol Rep. 2011 Dec;13(6):544–52

17. Wilson PW et al. “High density lipoprotein, low density lipoprotein and coronary artery disease.” Am J Cardiol. 1990 Sep 4;66 (6):7A–10A

18. Manoj Sigdel et al. “Association of High Sensitivity C-Reactive Protein with the Components of Metabolic Syndrome in Diabetic and Non-Diabetic Individuals”. J Clin Diagn Res. 2014 Jun; 8(6): CC11–CC13.

19. Shelbaya S et al. “Study of the role of Interleukin-6 and high sensitivity C-reactive protein in diabetic nephropathy in type 1 diabetic patients.” Eur Rev Med Pharmacol Sci. 2012; 16 (2):176–82

20. Navarro JF, Mora C, Maca M, Garca J. Inflammatory parameters are independently associated with urinary albumin in type 2 diabetes mellitus. Am J Kidney Dis. 2003;42:53–61.

21. Pannacciulli N, Cantatore FP, Minenna A, Bellacicco M, Giorgino R, De Pergola G. Urinary albumin excretion is independently associated with C-reactive protein levels in overweight and obese nondiabetic premenopausal women. J Intern Med. 2001;250:502–7.

22. Gomes MB, Nogueira VG. Acute-phase proteins and microalbuminuria among patients with type 2 diabetes. Diabetes Res Clin Pract. 2004;66:31–9.

23. Hansen TK, Gall MA, Tarnow L, et al. Mannose-binding lectin and mortality in type 2 diabetes. Arch Intern Med. 2006;166:2007–1

24. Safiullah Amanullah et al. “Association of hs-CRP with Diabetic and Non-diabetic individuals.” Jordan Journal of Biological Sciences, Volume 3, Number 1, January 2010, ISSN 1995-6673,Pages 7–12

25. Shelbaya S et al. “Study of the role of Interleukin-6 and high sensitivity C-reactive protein in diabetic nephropathy in type 1 diabetic patients.” Eur Rev Med Pharmacol Sci. 2012; 16 (2):176–82

26. Holm J, Ravn J, Ingemann Hansen S. Urinary excretion of alpha1-microglobulin and albumin in acute myocardial infarction. Correlation with plasma concentrations of troponin I and C-reactive protein.” Scand J Urol Nephrol.2006;40:339–44

27. Lee WY, Park JS, Noh SY, et al. C-reactive protein concentrations are related to insulin resistance and metabolic syndrome as defined by the ATP III report. Int J Cardiol. 2004;97:101–6.

28. Currie G et al. “Proteinuria and its relation to cardiovascular disease.” Int J Nephrol Renovasc Dis. 2013 Dec 21;7:13–24

29. Agrawal et al. “Cardiovascular implications of proteinuria: an indicator of chronic kidney disease.” Nat Rev Cardiol. 2009 Apr ;6(4):301–11

30. Chu NV, Kong AP, Kim DD, Armstrong D, Baxi S, Deutsch R, Caulfield M, Mudaliar SR, Reitz R, Henry RR, Reaven PD. Differential effects of metformin and troglitazone on cardio-vascular risk factors in patients with type 2 diabetes. Diabetes Care. 2002;25(3):542–9.

